# An integrative method for COVID-19 patients’ classification from chest X-ray using deep learning network with image visibility graph as feature extractor

**DOI:** 10.1101/2021.11.17.21266472

**Authors:** Mayukha Pal, Yash Tiwari, T. Vineeth Reddy, P. Sai Ram Aditya, Prasanta K. Panigrahi

**Affiliations:** ABB Ability Innovation Center, Asea Brown Boveri Company, Hyderabad 500084, India; Department of Physics, Indian Institute of Technology Hyderabad, Kandi, Sangareddy, Telangana 502285, India; Department of Mechanical and Aerospace Engineering, Indian Institute of Technology Hyderabad, Kandi, Sangareddy, Telangana 502285, India; Department of Artificial Intelligence, Indian Institute of Technology Hyderabad, Kandi, Sangareddy, Telangana 502285, India; Indian Institute of Science Education and Research Kolkata, Mohanpur 741246, India

**Keywords:** COVID-19 and SARS Coronavirus, Chest X-ray and Image classification, Haar wavelet and Image Visibility Graph, Assortative coefficient and Classification learners, Resnet34 and Convolutional neural network, Multilayer perceptron classifier

## Abstract

We propose a method by integrating image visibility graph and deep neural network (DL) for classifying COVID-19 patients from their chest X-ray images. The computed assortative coefficient from each image horizonal visibility graph (IHVG) is utilized as a physical parameter feature extractor to improve the accuracy of our image classifier based on Resnet34 convolutional neural network (CNN). We choose the most optimized recently used CNN deep learning model, Resnet34 for training the pre-processed chest X-ray images of COVID-19 and healthy individuals. Independently, the preprocessed X-ray images are passed through a 2D Haar wavelet filter that decomposes the image up to 3 labels and returns the approximation coefficients of the image which is used to obtain the horizontal visibility graph for each X-ray image of both healthy and COVID-19 cases. The corresponding assortative coefficients are computed for each IHVG and was subsequently used in random forest classifier whose output is integrated with Resnet34 output in a multi-layer perceptron to obtain the final improved prediction accuracy. We employed a multilayer perceptron to integrate the feature predictor from image visibility graph with Resnet34 to obtain the final image classification result for our proposed method. Our analysis employed much larger chest X-ray image dataset compared to previous used work. It is demonstrated that compared to Resnet34 alone our integrative method shows negligible false negative conditions along with improved accuracy in the classification of COVID-19 patients. Use of visibility graph in this model enhances its ability to extract various qualitative and quantitative complex network features for each image. Enables the possibility of building disease network model from COVID-19 images which is mostly unexplored. Our proposed method is found to be very effective and accurate in disease classification from images and is computationally faster as compared to the use of multimode CNN deep learning models, reported in recent research works.

**Significance:** An integrative method is proposed combining convolutional neural networks and 2D visibility graphs through a multilayer perceptron, for effective classification of COVID-19 patients from the chest x-ray images. In our study, the computed assortative coefficient from the horizontal visibility graph of each wavelet filtered X-ray image is used as a physical feature extractor. We demonstrate that compared to Resnet34 alone, our proposed integrative approach shows significant reduction in false negative conditions and higher accuracy in the classification of COVID-19 patients. The method is computationally faster and with the use of visibility graph, it also enables one to extract complex network based qualitative and quantitative parameters for each subject for additional understandings like disease network model building and its structures etc.

## 1. Introduction

SARS-CoV-2 virus is highly infectious and spreads faster, affecting the respiratory organs like lungs, with the respiratory tracks developing various breath related symptoms in the patients. The severity of the disease in the humans is based on the spread of the infection to the respiratory organs. Patients suffer with heavy cough, high fever, muscle/body pain, sore throat, loss of sensation for taste and smell, headache, fatigue, and shortness of breath. In case of severe infection, the patient’s oxygen saturation level drops drastically bringing more medical complications requiring immediate oxygen support and/or intensive medical care. The disease is named as COVID-19 by the World Health Organization which declared it as a Pandemic [1-3]. Across the globe, it is observed that proper social distancing, wearing of masks covering nose and mouth, and proper sanitization effectively controls the spread. It is also consistently observed that breaking the chain of the infection by imposing lockdown restricting human movement and imposing COVID-19 containment protocol effectively controls the spread in case of an infection outbreak [4].

Continuous lockdown adversely affects the economic activities and GDP of a nation. It also severely impacts the livelihood of the population working as daily wage laborers, in unorganized sectors, self-employment businesses and for the citizens below poverty line. Hence it is essential to balance the economic activities ensuring all sections of the society are able to sustain their lives, while the nation effectively manages and controls the virus spread. More scientific approaches like early detection, test automation and other tools would help administration effectively manage and control the situation [5-6]. Various nations have also started vaccination programs to increase the human body immunity to fight against the coronavirus and its mutant strain variants. Another approach of effectively fighting against the virus is to test and diagnose the disease early so that self-isolation is maintained to further stop the virus spread and also effective medication is started early for the patient to stop the infection spread within the body [7]. Nearly 251 million people across the globe have been infected with the coronavirus with more than 5 million deaths. During surge in the virus inspection, healthcare infrastructures face acute shortage of medicines, radiology test facilities, medical oxygen, medical equipment and ICU beds to cater the surge in the high demand challenging test and hospital facility. Generally rapid antigen test (RAT), reverse transcription-polymerase chain reaction (RT-PCR) tests are performed for initial diagnosis of COVID-19. Many instances these tests showed false negatives hence are not much reliable [8-10]. Hence clinicians prefer chest radiology tests to check the lungs image for the COVID-19 Pneumonia. As computed tomography (CT) scans subject more radiation exposure for the patients with limited available facilities compared to X-ray hence X-ray is preferred for initial investigation. For a mass scale community infection, chest X-ray is a good, low-cost, first-look diagnostic tool with quick results. Also, availability of handheld X-ray devices allows easy access and early diagnosis in rural areas hence improves test penetration for effective control of infection spread through early detection.

Various machine learning and deep learning tools are used for classification of COVID-19 patients from their chest X-ray images like Random Forest, Support Vector Machine, Artificial Neural Networks (ANN), convolutional neural network (CNN) [11-20]. Generally, accuracy of various deep learning methods varies from 78-98% for classifying the COVID-19 patients based on their X-ray images [21-31]. Recently proposed MSSIM based method with the use of a simple classification learner showed 97.7 % accuracy for detecting COVID-19 X-ray images with lower false negative case [32]. As the method is comparing between two images to quantify the differences hence generates large data points due to cross-correlation analysis while using the same given dataset, making robust and accurate training for the classification learner. In the present study, we aim to use X-ray images of the patients to find characteristic features for classification using DL without use of comparison-based analysis as used in MSSIM method. Inspired by this, we propose here a new method by integrating CNN and visibility graph for faster computation and accurate classification with low false negative compared to currently used deep learning algorithms for identification of COVID-19 patients from X-ray image. Our choice of Resnet34 is due to the fact that, it is the most optimized and recently analyzed deep convolutional neural networks [33-34] that is used for image classification, object detection, image segmentation applications as it resolves the inherent problem of degrading gradient descent through residual blocks adding the skip connections in the identity mapping. The section 2 of the manuscript details the chest x-ray data and the methods used in the analysis while section 3 of the manuscript discusses the results from our analysis. Section 4 of the manuscript gives our conclusion and inference to the work.

## 2. Materials and Methods

The COVID-19 X-ray image database [35] is used to obtain 500 X-ray images of patients diagnosed with COVID-19. 500 healthy subject’s Chest X-rays is obtained from the open-source database [36]. These pre-processed images are used for our CNN analysis. Further for our visibility graph analysis, these X-ray images are resized to make them of equal pixel size after converting to grayscale first. The images were then resized to 1024×1024 and these image matrices are of data type double. We applied 3 label Haar wavelet decomposition and considered the approximation coefficients for our visibility graph analysis. Here to compress the image, we used 2D Haar discrete wavelet transform (HDWT). Generally, Haar wavelet transform is used to perform lossy image compression to ensure the compressed image retains its quality. This is one of the efficient procedures to perform lossless and lossy image compression as it uses averaging and differencing values in an image matrix to produce a matrix which is mostly sparse having less non-zero element in the matrix [37]. Haar wavelet transform utilizes a rectangular window for sampling. In the first label decomposition, a window width of two is used and the width doubles at each step until the window encompasses the entire data on hand. Each decomposition generates a new time series and a set of coefficients where the new time series is the average of the previous label time series over the sampling window and the coefficients represent the average change in the sampling window. Let us assume a time series {x_i_, x_i+1_, x_i+2_,…} then the Haar wavelet coefficient is defined as: 

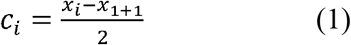

The Haar scaling function is written as

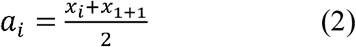

The Haar scaling function gives the average version of the data set and is half the size of the input time series. As the analysis procedure is recursive, the average or the smoothed data becomes the input for the next label of the wavelet transform. Unlike other wavelet functions, in Haar wavelet transform, it preserves the average in the smoothed values. The obtained average coefficient from Haar wavelet in our visibility graph analysis contains all information about the image and reduces matrix dimension to 128×128 size allowing extreme fast computation by reducing months of time required in the computation for such image visibility graph analysis to a few hours. Fig. 1 shows X-ray images of a COVID-19 subject obtained from the database along with a healthy chest image. Fig. 2 represents the sample plot of the approximation coefficient obtained from 2D Haar wavelet for COVID-19 and healthy subjects.

**Figure-1:**
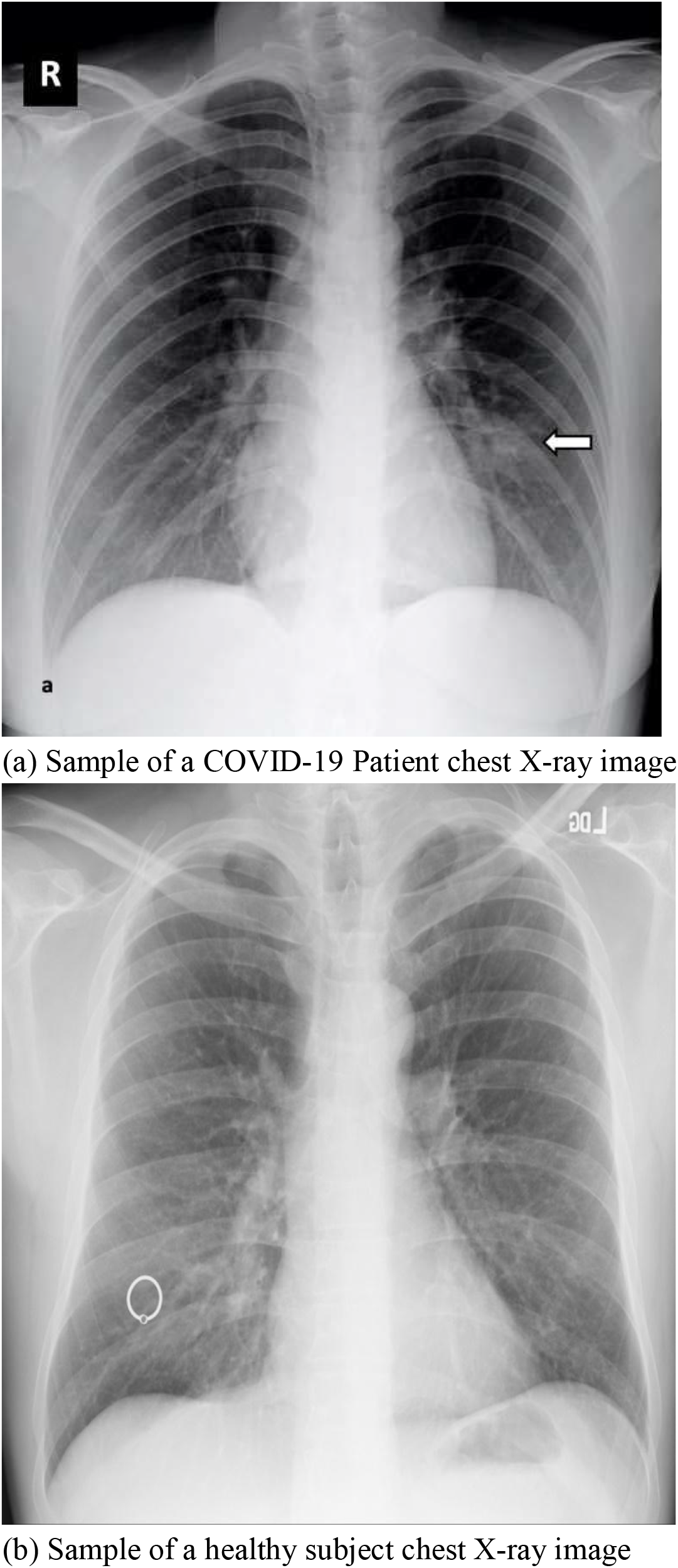
Sample chest X-ray images of different subject types.

**Figure-2:**
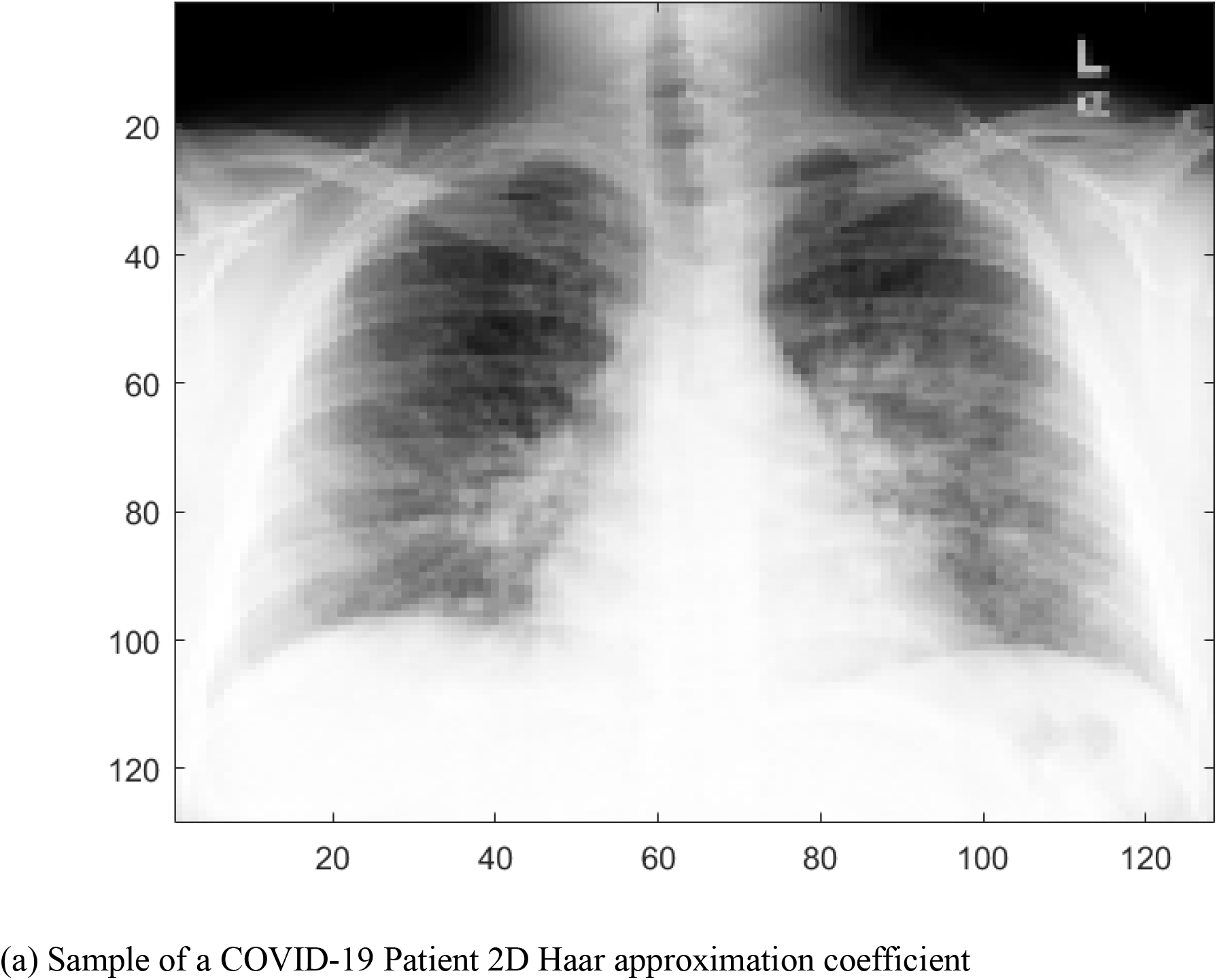
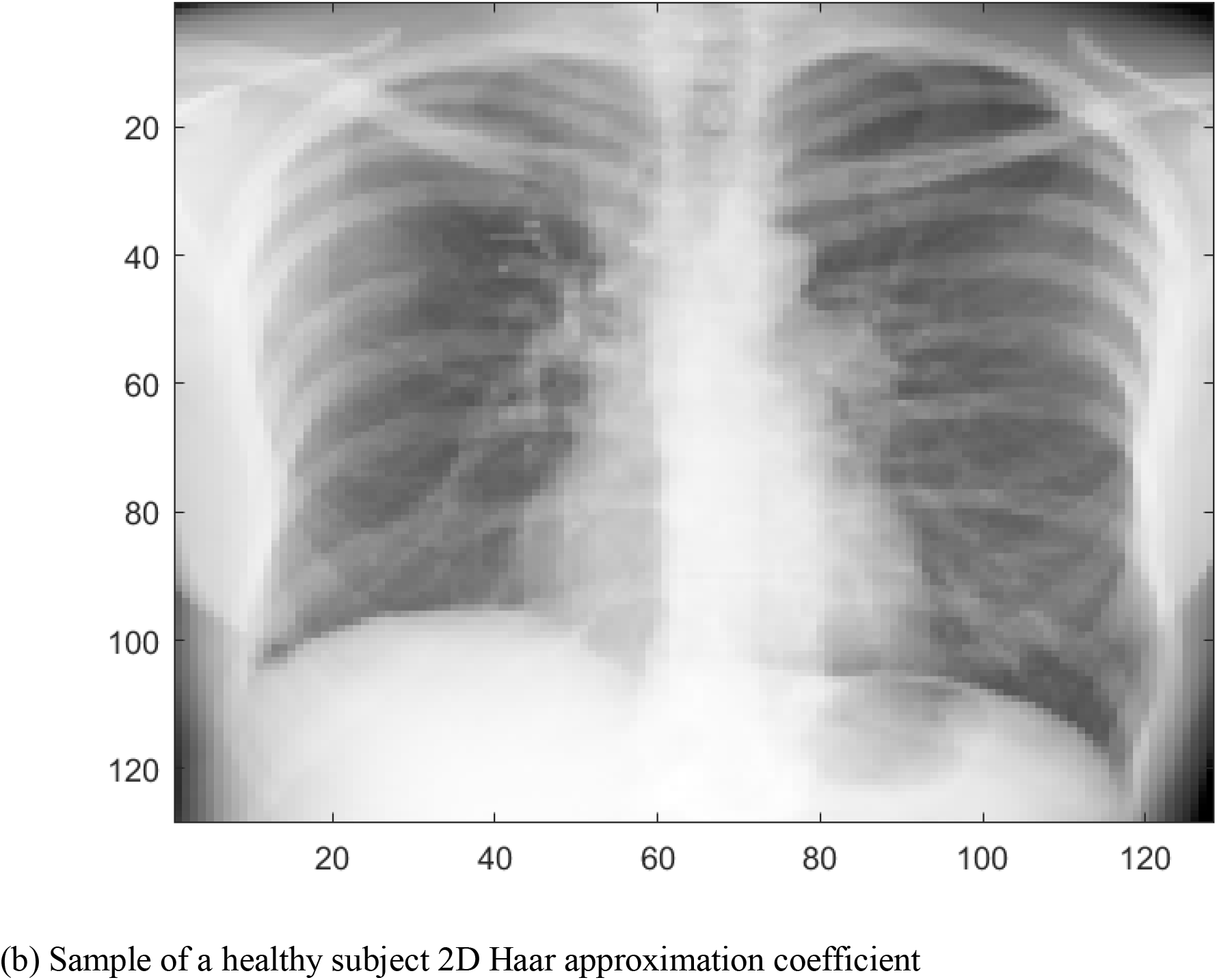
Sample chest X-ray images from the 2D Haar approximation coefficient after label 3 decomposition of the images.

Further these obtained approximation coefficient matrices from Haar wavelet for each image are considered in image visibility graph analysis [38]. If the analyzing matrix is a NxN for the X-ray image I where I_ij_ Є □ then the image visibility graph (IVG) will have N^2^ nodes. Now each node can be labelled by the indices of its corresponding datum I_ij_ in a manner that two nodes ij and i’j’ are linked if (i = i’) V (j = j’) V [(i = i’ +p)□(j = j’ +p)], for some integer p, and if I_ij_ and I_i’j’_ are linked in the visibility graph defined over the ordered sequence which includes ij and i’j’. We define the visibility graph (VG) as an undirected graph of n nodes, where each node i is labelled according to the time order of its corresponding datum x_i_. For visibility graph, two nodes i and j where i < j are connected by an undirected link if and only if a straight line connecting x_i_ and x_j_ can be drawn without intersecting any intermediate datum x_k_ for i < k < j and also the convexity criterion is fulfilled: x_k_ < x_i_ + ((k-i)/(j-i)) [x_j_ - x_i_]; □k : i < k < j. For horizontal visibility graph (HVG) the two nodes i, j where i < j is connected by a link if and only if a horizontal line can be drawn connecting x_i_ and x_j_ such that it does not intersect any intermediate datum x_k_ for i < k < j. Also, we connect i and j in the HVG if ordering criterion is fulfilled: x_k_ < inf(x_i_, x_j_), □k : i < k < j. The image horizontal visibility graph (IHVG) obeys the same set of conditions like IVG [39-41].

We computed graphs from the coefficient matrices of the images using IHVG, where pixels are considered as nodes and the nodes are connected if they lie in a specific direction i.e. rows, columns, diagonals as per the defined HVG visibility criteria. We obtained 500 IHVG graphs for COVID-19 patients and 500 for the healthy subjects. Then from each IHVG graph the assortative co-efficient is computed as a physical feature extractor and used along with our earlier obtained Resnet34 results through a multilayer perceptron (MLP) to obtain the final classification results. As the graph for each image is available, we could obtain qualitative and quantitative characteristics like degree distribution, image descriptor patch frequency, assortative coefficient etc from the graph for characterization studies and further for possible disease network model development.

In our Resnet34 model, we normalized the pixel values of the input images using mean values [0.485, 0.456, 0.406] and the standard deviation values [0.229, 0.224, 0.225] then further the values were rescaled in range of -2 to 2. In CNN, image normalization is employed for faster convergence, easy training hence faster learning speed with stable gradient descent. Image augmentation is used in CNN model to improve the network generalization ability however we didn’t use in Resnet34 model to avoid data bias due to its use in our integrative model with image visibility graph.

We used random forest classifier for the assortative coefficients obtained from visibility graph and integrating with Resnet34 results through MLP. The classifier was chosen based on the performance from host of different classifier learners. Among various CNN models analyzed like Resnet18, Resnet50, we found Resnet34 to be performing better. We used 224×224×3 input layers with 50 epochs in our Resnet 34 model. We modified the bottom fully connected (fc) layer of our Resnet34 model with a two-neuron fc layer followed by a softmax activation output layer for computing the probabilities. The learning rate of the Resnet34 model is set to 0.001 and uses cross-entropy validation. For integrating the Resnet34 with visibility graph, we made use of an MLP classifier having relu activation function, adam optimizer with 100 hidden layers for maximum iteration of 500.

## 3. Results and Discussion

From the visibility graph analysis, IHVG graphs were obtained for all preprocessed image matrices. A sample IHVG graph is shown in Fig 3. To understand various qualitative and quantitative properties of these graphs, we counted the repetitions of small subgraphs in the IHVG associated to a given input image data matrix which is referred to as visibility patches (VPs). The computed patch frequency is a local image property descriptor. Visibility patch of order p is defined as, VP_p_ which is any subgraph from the IHVG formed by a set of p^2^ nodes 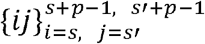 for arbitrary s,s’ satisfying the condition 1≤s, s’ ≤N-p. N equal to 128 is the size of NxN data matrix in our visibility graph analysis. In the analysis, lowest order p=3 yields nontrivial visibility patches. Visibility patches are detected when sliding a 3×3 pixel cell of stride 1 in the image extracting the corresponding IHVGs within the cell. It enables reduced checking of different combination of visibility graph motifs presence or absence hence mathematically enables tractability of visibility patches for computation. Fig 4 shows the visibility patches for 3 randomly selected X-ray images of both healthy and COVID-19 subjects.

**Figure-3:**
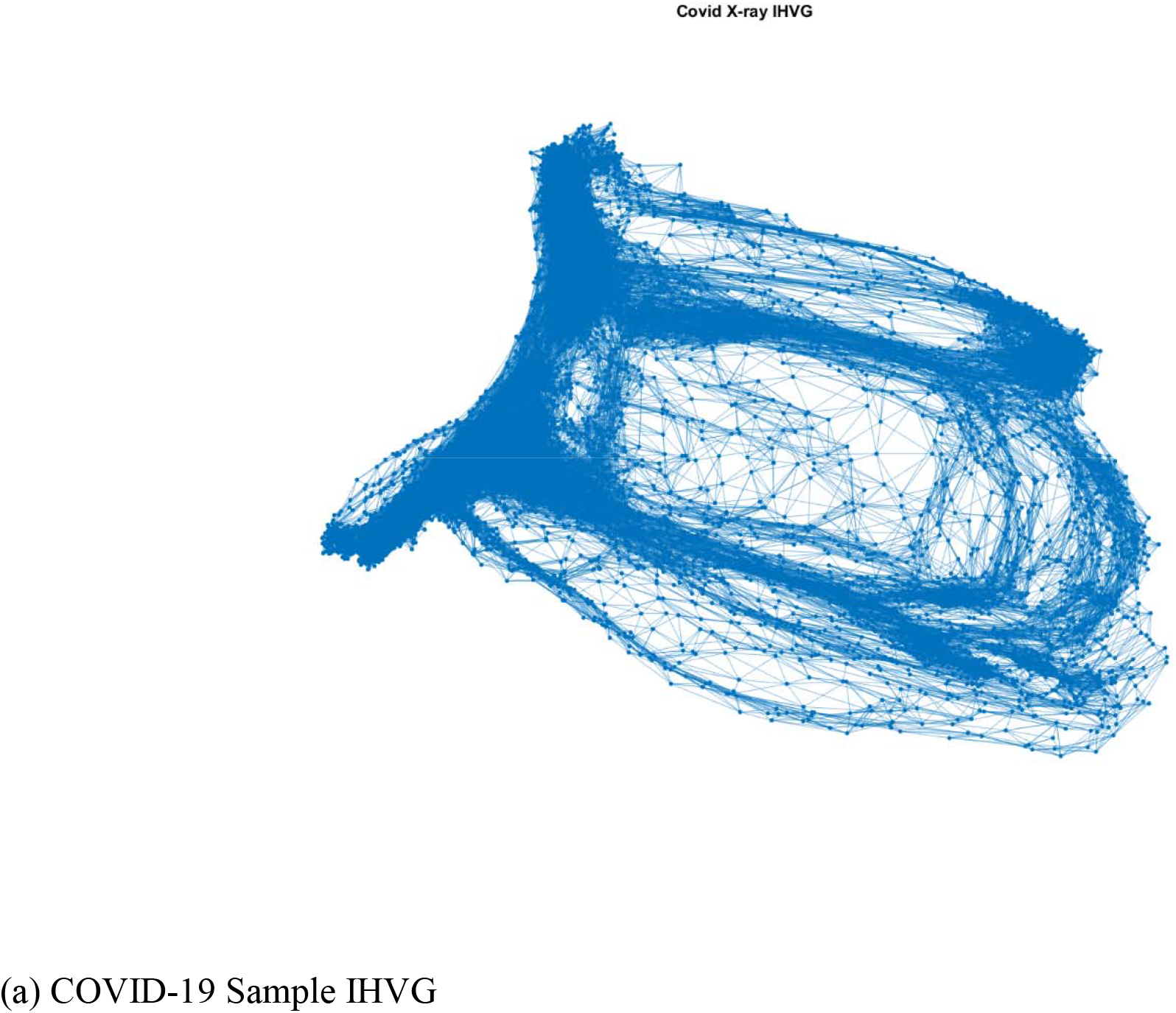
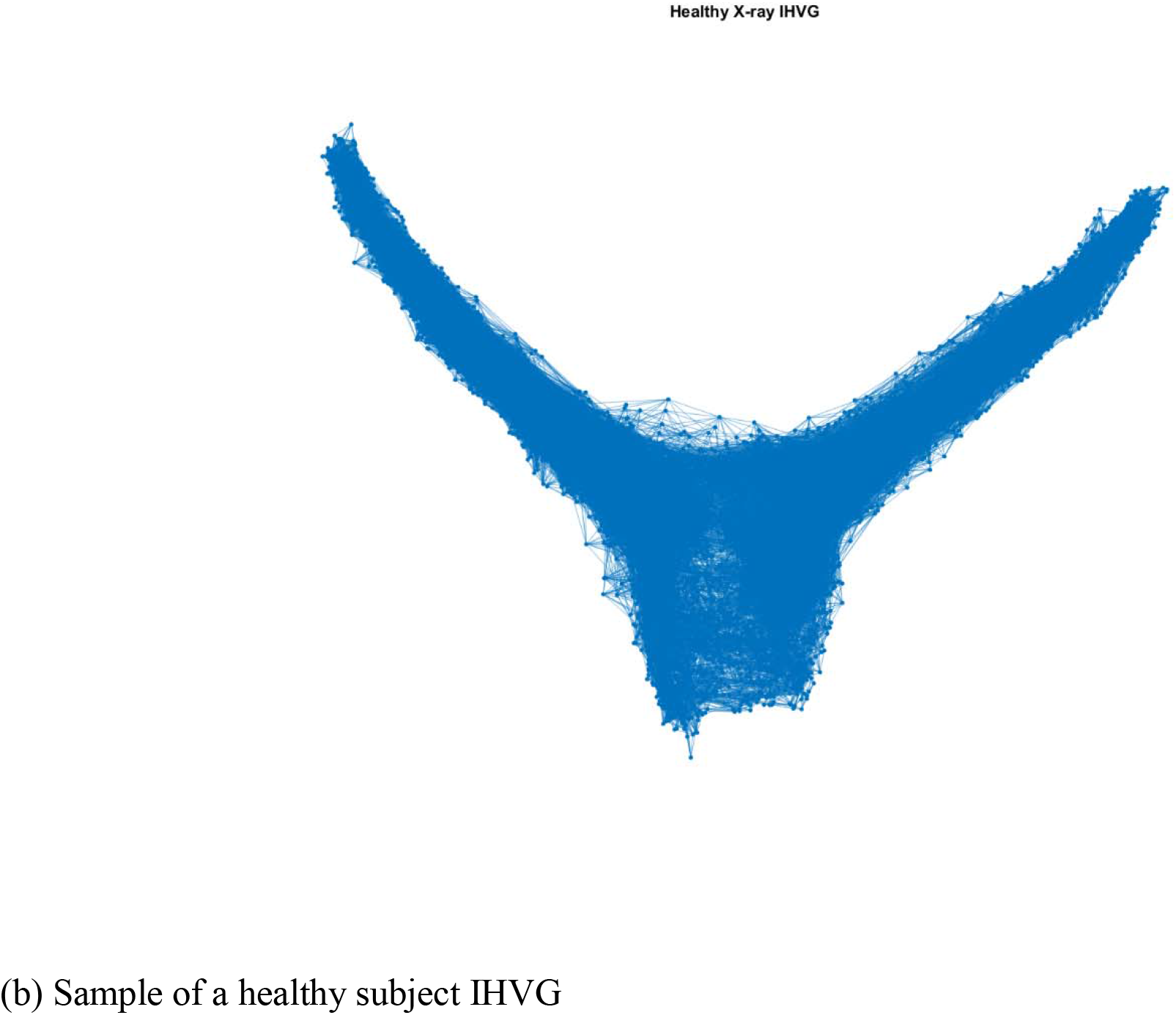
A Sample IHVG graphs for a COVID-19 patient and a healthy subject.

**Figure-4:**
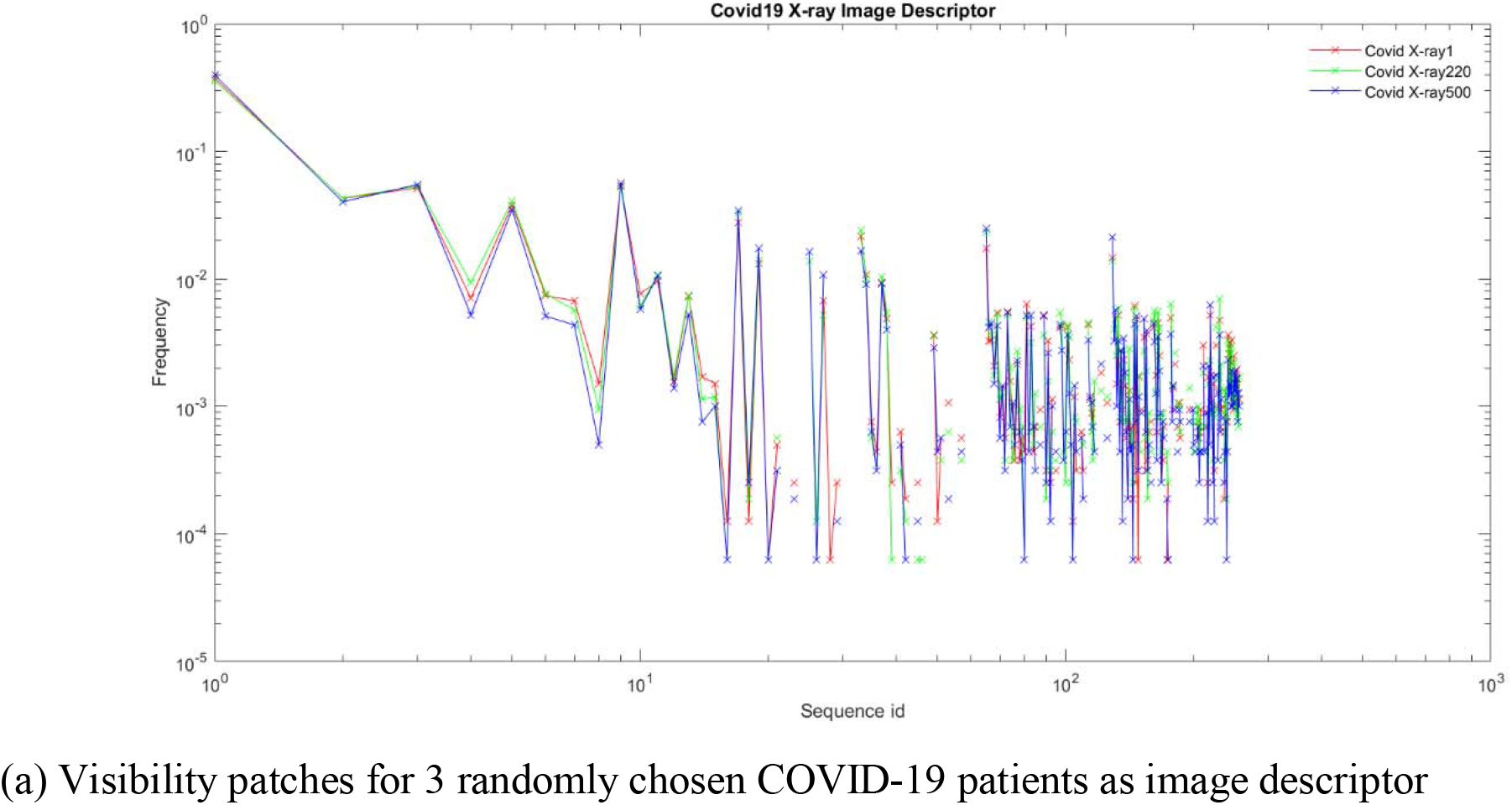
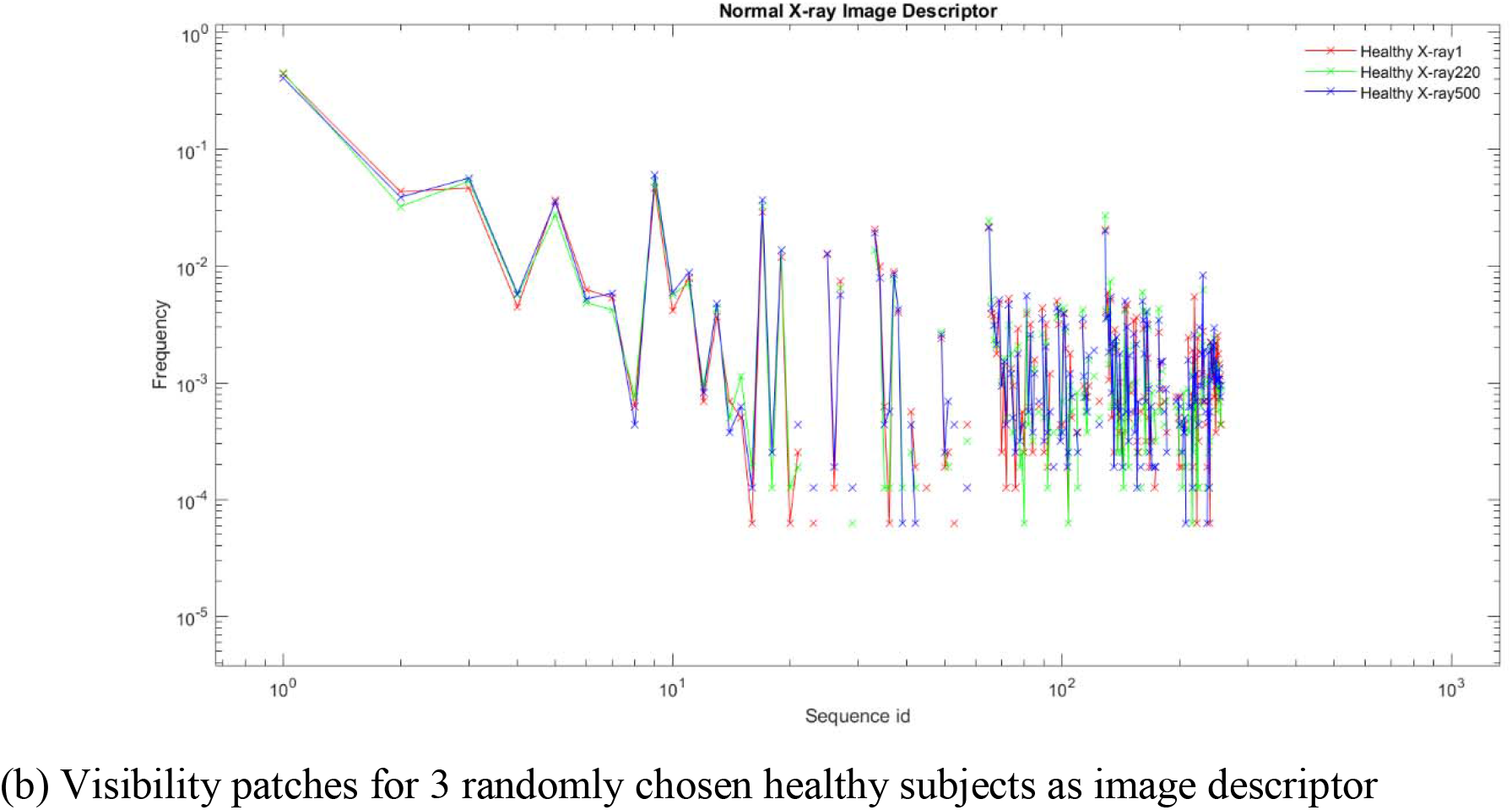
Visibility patches for both COVID-19 and healthy subjects.

We analyzed the degree distribution P(k) where P(k) ∼ k^-λ^ to understand statistical properties of the networks where k is the degree of a node and is defined as the number of edges connected to a node. If the degree distribution of the network, follow a power law distribution then we call it a scale-free network which is characterized by the nodes of the networks that are linked to a significant fraction of the total number of edges of the network. Log-log plot of the degree distributions P(k) for a sample IHVG is shown in Fig 5. We observe that for both healthy and COVID-19 patients the obtained network is scale free.

**Figure-5:**
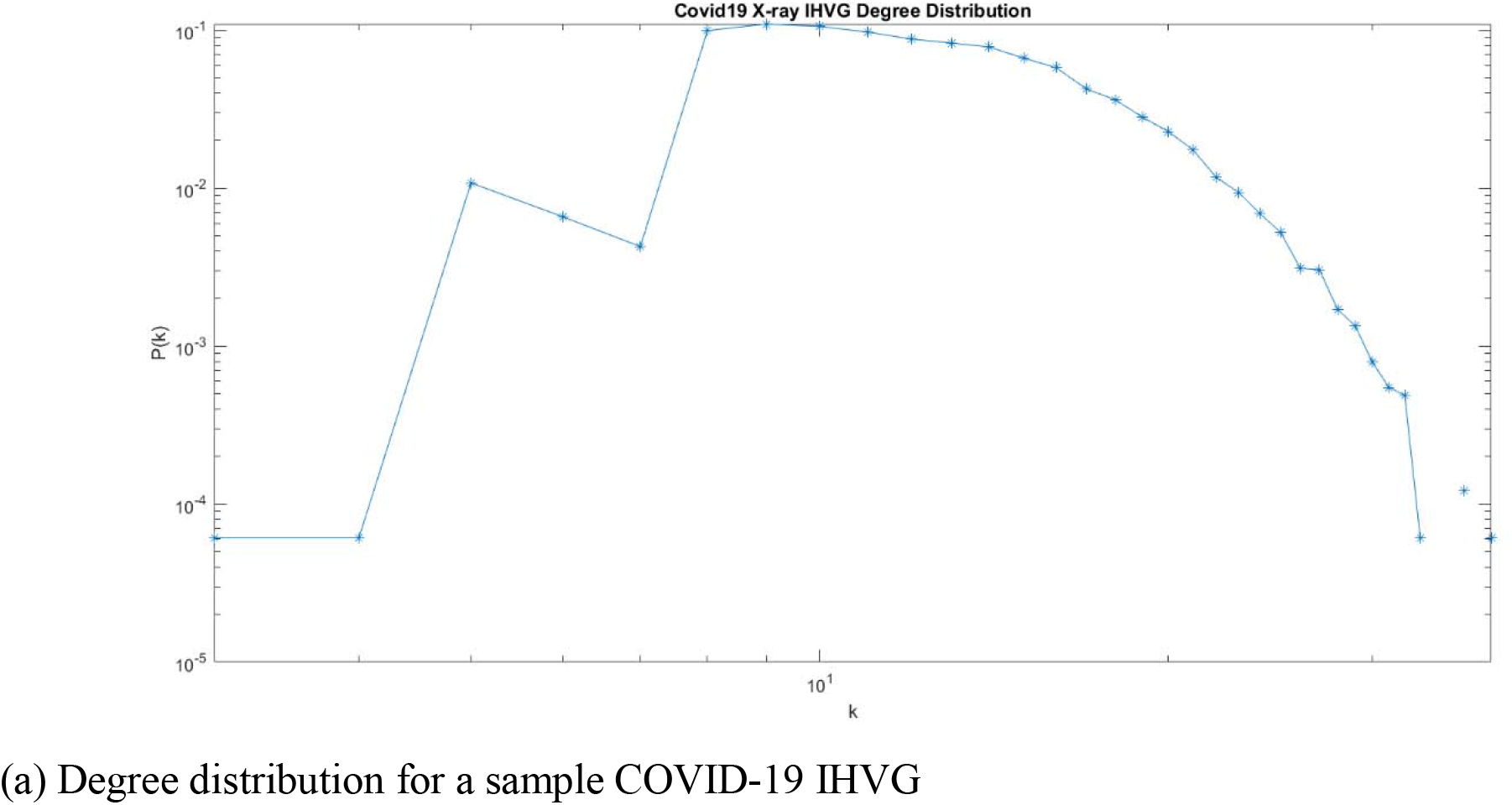
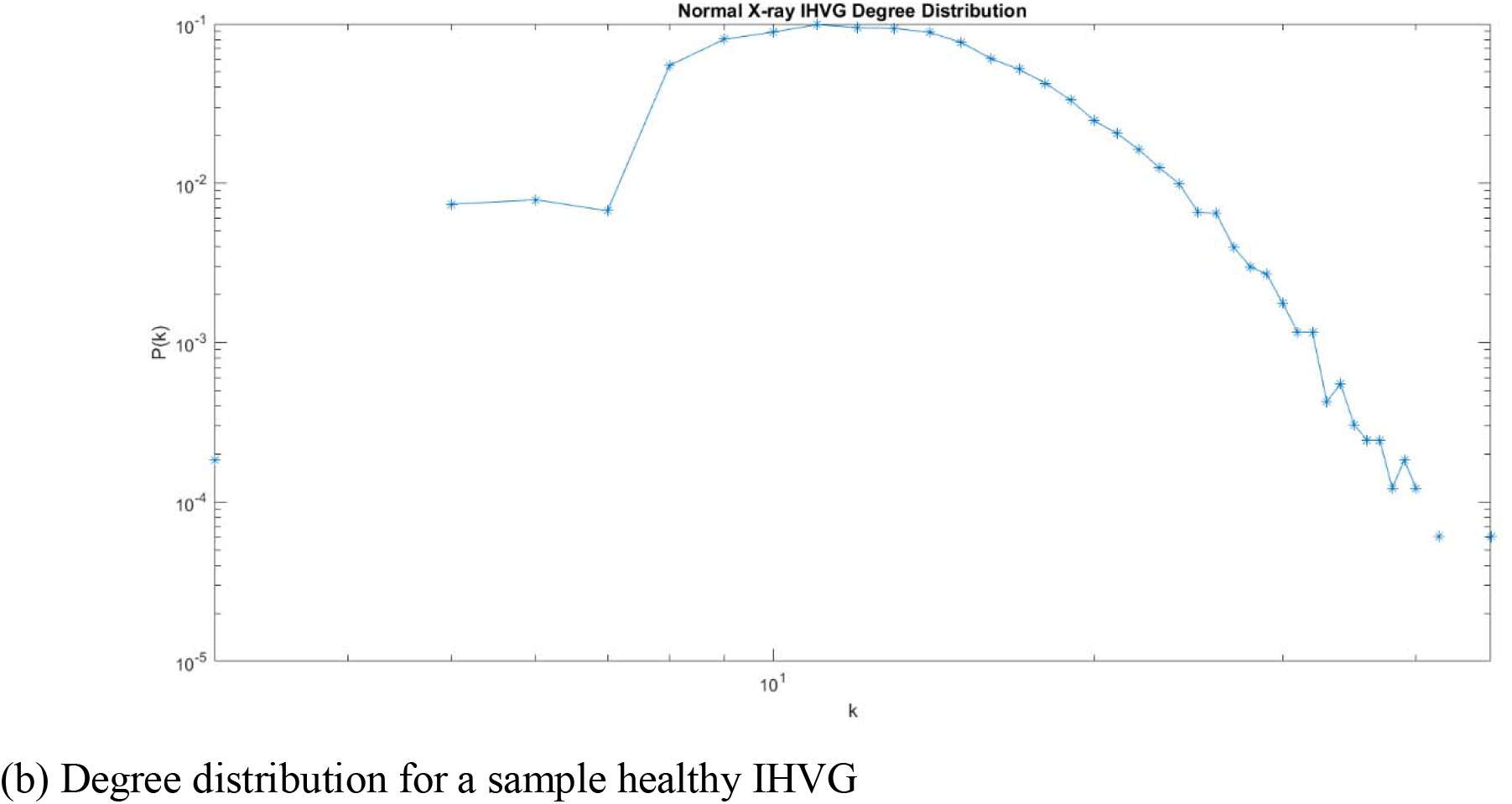
The degree distribution from the IHVG graph for sample COVID-19 and healthy subjects.

We computed assortative coefficients for each obtained IHVG which is used along with Resnet34. A complex network is defined as assortative mixing if higher degree nodes of the network tend to be connected to other higher degree nodes whereas in disassortative mixing the high degree nodes attach to low degree nodes only. Computationally, these assortative complex networks remove its highest degree nodes efficiently compared to the disassortative networks. For an undirected network, the assortative coefficient is computed as:

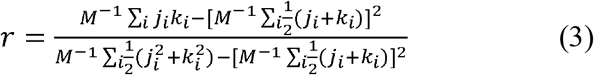

where j_i_, k_i_ are the degrees of the two nodes at the ends of the i^th^ edge where i=1,2, upto M. Assortative coefficient values vary between -1 to 1 where assortative networks have positive r values while disassortative networks have negative r values. For both COVID-19 and healthy subjects in our analysis, the obtained assortative coefficient values are always positive indicating that all the networks for disease and healthy are assortative.

The obtained 1000 assortative coefficients for both COVID-19 and healthy are labeled with its attribute category for random forest classifier and are further used along with Resnet34 in a MLP classifier as a quantitative physical parameter feature extractor for improving classification performance of standalone Resnet34 as visualized from the confusion matrix. Fig 6 illustrates our system study approach.

**Figure-6:**
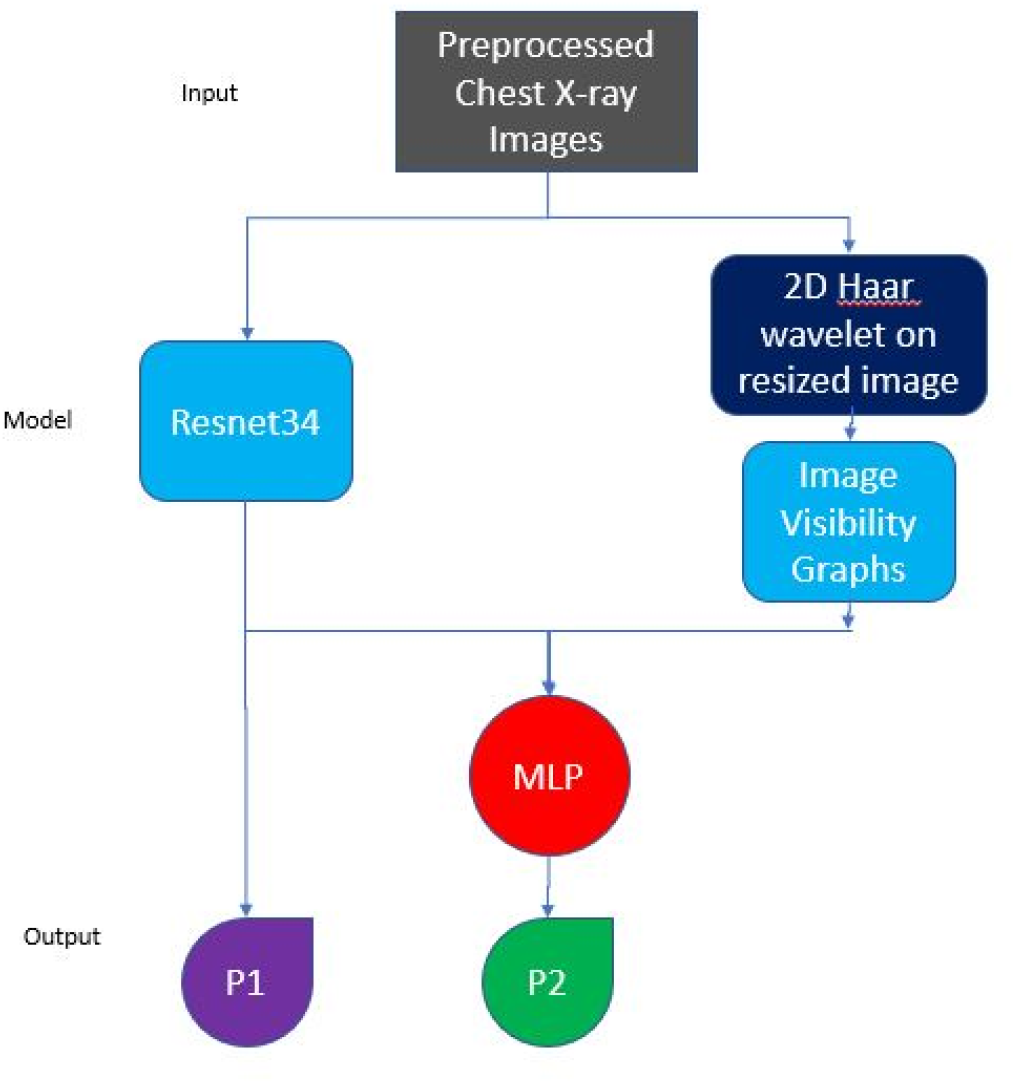
Our proposed integrative method in analyzing COVID-19 X-ray images for classification.

It is worth emphasizing that when the sample size of chest X-ray images increases as in our case to 1000 images, the resnet34 model without image augmentation gives 96% classification accuracy unlike earlier reported accuracy of 98.33% with 406 images [33]. We avoided image augmentation in our CNN model to overcome the data bias error that might occur in testing. As shown in Fig6, we make use of Resnet34 on the X-ray images to classify the COVID-19 and healthy subjects. In a parallel approach, from the preprocessed images passing through Haar wavelet visibility graph was introduced to learn the structural connectivity from the obtained network and then random forest classifier is used on VG assortative coefficient to classify the images. Then we integrate both Resnet34 and VG using a MLP considering the predicted labels from both for the final classification result of MLP. In our model, we used 1000 chest X-ray images of which 500 were for COVID-19 while 500 of healthy. We used 800 of these images equally from both the class for training the Resnet34 model and the random forest classifier using assortative co-efficient from visibility graph. The remaining 200 is used for validation. The predicted labels from random forest and Resnet34 were fed to the MLP classifier in which 100 data points were used for training while 100 were used for testing to obtain the final accuracy of our model. Table 1 summaries performance of our proposed multi-mode ensemble model with recently developed models. We observe our proposed multi-mode ensemble with introduction of VG as a unique characteristic feature that increases the accuracy by 2% compared to use of Resnet34 model alone. Similarly, the F1-score, sensitivity is increased by almost 1.8% compared to the Resnet34 model alone. Importantly, from the confusion matrix as shown in Table 2 we also observe our integrative model decreases false negative cases proportionately. The Fig 7 shows our proposed method training and test accuracy and loss. Our integrative model loss against epoch is very close to 0 while it is 0.4 for Resnet34. The Fig 8 shows the receiver operating characteristic (RoC) curve for our newly proposed model.

**Table 1:** Comparison of our proposed method with state-of-the-art deep learning COVID-19 detection approaches using chest X-ray (CXR) images Confusion matrix for Resnet34 alone (P1)

| **Article Reference** | **Classification Method** | **Used Class** | **Number of CXR Images** | **Accuracy (%)** | **COVID-19 class Sensitivity (%)** |
| --- | --- | --- | --- | --- | --- |
| **Ref [13]** | COVIDX-Net | COVID-19<br>Normal | Total – 50<br>COVID-19: 25<br>and Normal: 25 | 90.00 | 100.00 |
| **Ref [42]** | ResNet-50<br>and SVM | COVID-19<br>Normal | Total – 50<br>COVID-19: 25<br>and Normal: 25 | 95.38 | NA |
| **Ref [16]** | ResNet-50 | COVID-19<br>Normal | Total – 100<br>COVID-19: 50<br>and Normal: 50 | 98.00 | 96.00 |
| **Ref [43]** | SqueezeNet<br>and<br>MobileNetV2<br>SMO and<br>SVM | COVID-19<br>Normal<br>Pneumonia | Total – 458<br>COVID-19: 295<br>Normal: 65 and<br>Pneumonia: 98 | 98.25 | 99.32 |
| **Ref [14]** | COVID-Net | COVID-19<br>Normal<br>Pneumonia | Total – 13800<br>COVID-19: 183<br>Normal: NA and<br>Pneumonia: NA | 92.60 | 87.10 |
| **Ref [44]** | Bayes-SqueezeNet | COVID-19<br>Normal<br>Pneumonia | Total – 5949<br>COVID-19: 76<br>Normal: 1583<br>and<br>Pneumonia: 4290 | 98.30 | 100.00 |
| **Ref [23]** | DarkCovidNet | COVID-19<br>Normal | Total – 625<br>COVID-19: 125<br>and Normal: 500 | 98.08 | 90.65 |
|  |  | COVID-19<br>Normal<br>Pneumonia | Total – 1125<br>COVID-19: 125<br>Normal: 500 and<br>Pneumonia: 500 | 87.02 |  |
| **Ref [33]** | ResNet-34 | COVID-19<br>Normal | Total – 406<br>COVID-19: 203<br>and Normal: 203 | 98.33 | 100.00 |
| **Ref [32]** | SSIM<br>Coefficient<br>with<br>Ensemble<br>Tree<br>Classifier | COVID-19<br>Normal | Total – 1000<br>COVID-19: 500<br>and Normal: 500 | 97.7 | 97.62 |
| **Proposed**<br>**Method** | Resnet34 with<br>Visibility<br>Graph | COVID-19<br>Normal | Total – 1000<br>COVID-19: 500<br>and Normal: 500 | 98 | 97.87 |

**Table-2:**
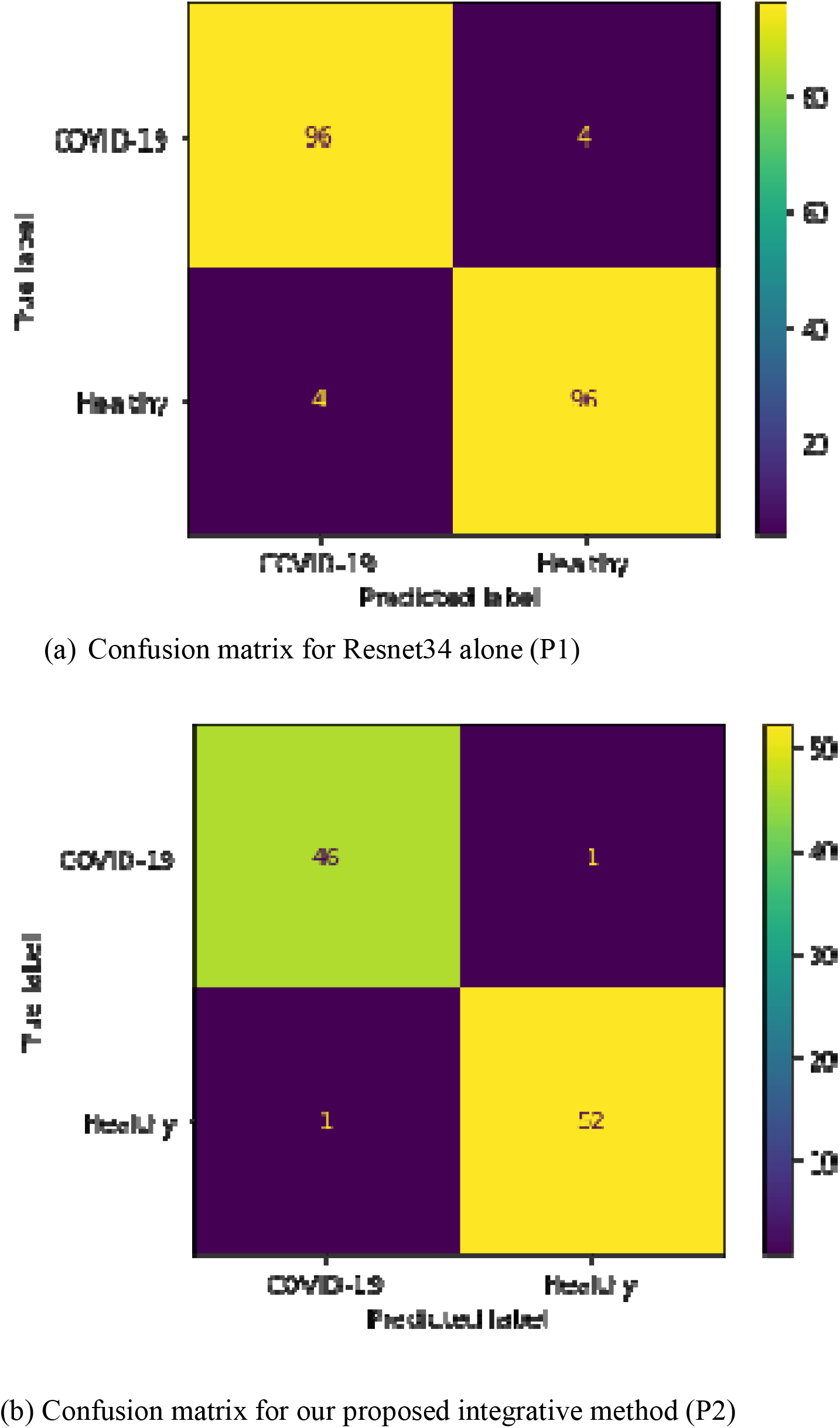
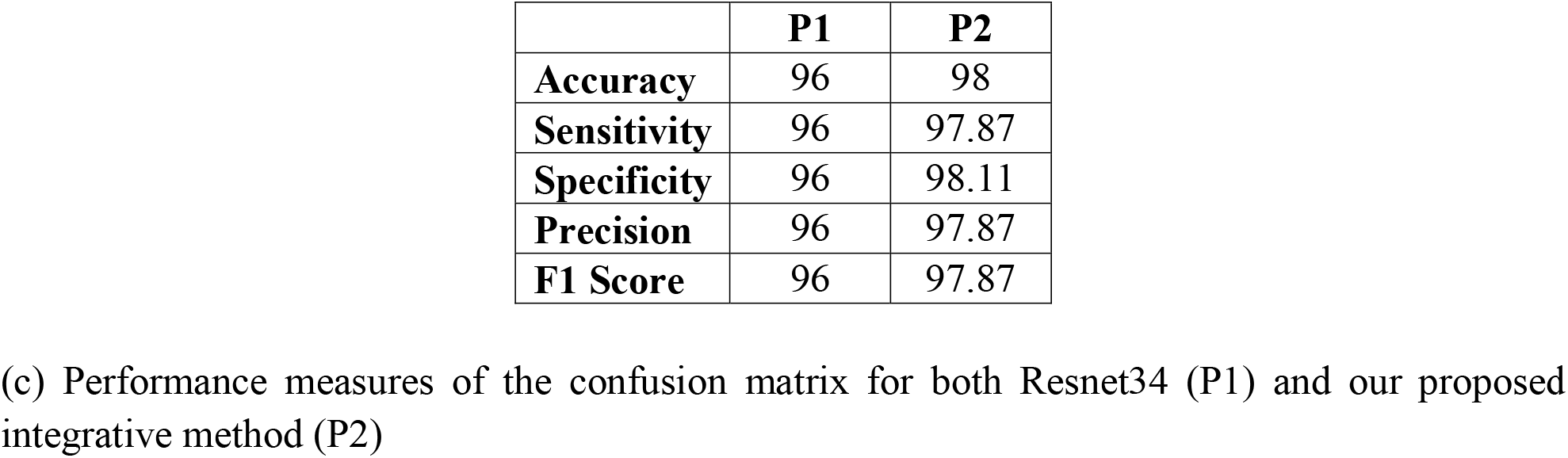
Confusion matrix for the classifiers with its performance measures.

**Figure-7:**
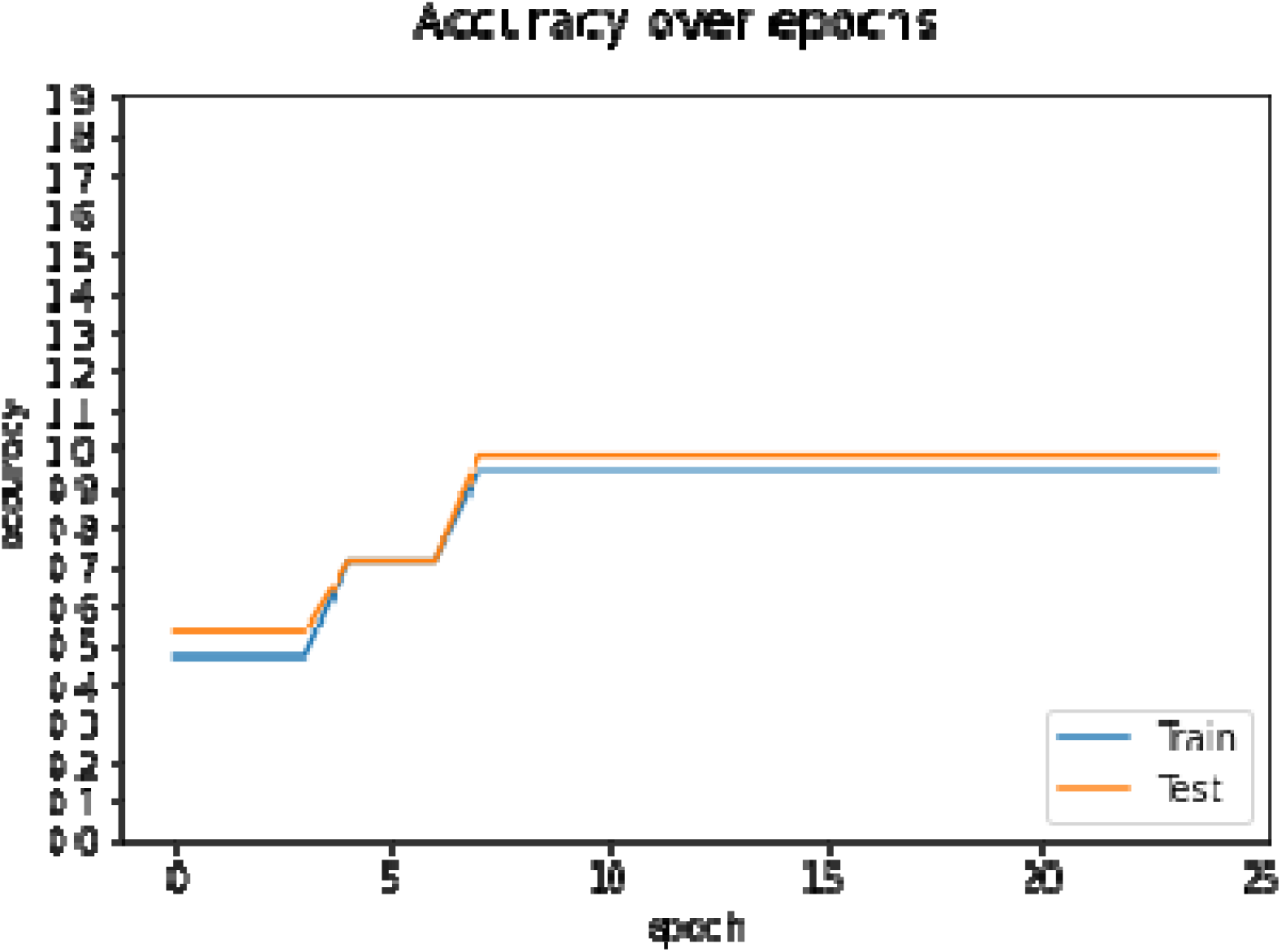
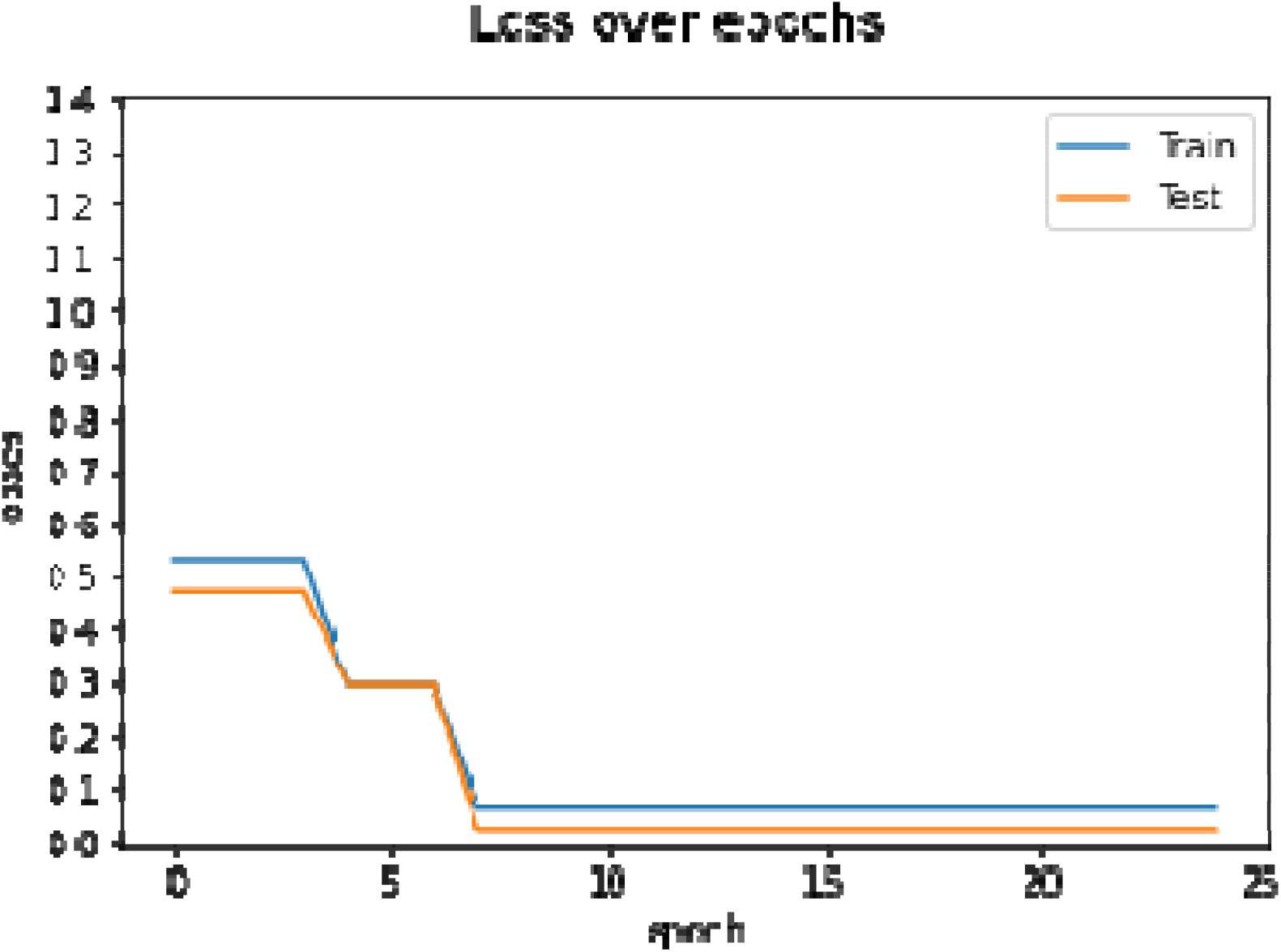
Our proposed integrative method training and test accuracy, loss over epochs

**Figure-8:**
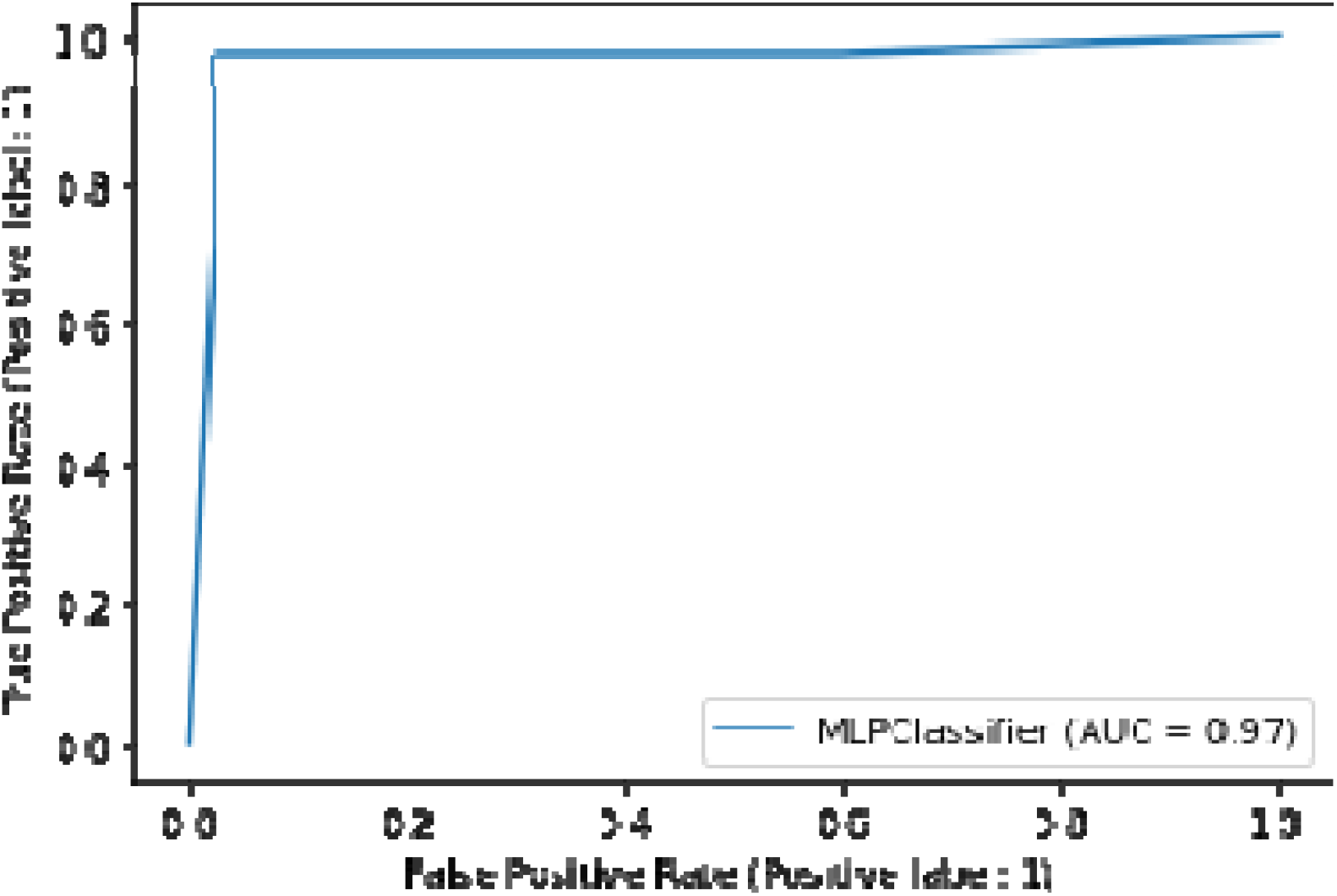
The RoC curve for our newly proposed model

## 4. Conclusion

In conclusion, we have shown our integrative multi-model ensemble method combining Resnet34 CNN model and 2D visibility graph helps in better classification of COVID-19 chest X-ray images compared to a CNN model alone or in combinations as reported in prior research work. Also, our method is computationally very fast and with introduction of Haar wavelet it drastically reduced computation time for image visibility graph and its associated parameter calculation while improving classification performance. With introduction of visibility graph various complex network qualitative and quantitative parameters for the subject image could be obtained and a disease network model could be built for future research on COVID-19.

## Data Availability

The COVID-19 X-ray image database [35] is used to obtain 500 X-ray images of patients diagnosed with COVID-19. 500 healthy subjects Chest X-rays is obtained from the open-source database [36].

https://github.com/ieee8023/COVID-chestxray-dataset.

## Acknowledgements

This research work did not receive any specific grant from funding agencies in the public, commercial, or not-for-profit sectors. The author alone is responsible for the content and writing of the paper.

## Authors’ contributions

MPal conceived the idea and conceptualized it, developed the model concept and its code, performed the data pre-processing and image processing analysis, wrote and reviewed the manuscript. YT performed the CNN and MLP related code enhancement, analysis, model optimization and contributed to the manuscript writing. TVR developed primary model framework and code. PSRA debugged the code and performed CNN analysis. PKP conceptualized the idea of using haar wavelet with visibility graph, mentored the work and reviewed the manuscript.

## Notes

### Competing Interest Statement

The authors have declared no competing interest.

### Author Declarations

The data obtained for our analysis is from the available public domain database made for academic research purpose and are appropriately cited in this work. The COVID-19 X-ray image database [35] is used to obtain 500 X-ray images of patients diagnosed with COVID-19. 500 healthy subjects Chest X-rays is obtained from the open-source database [36]. https://github.com/ieee8023/COVID-chestxray-dataset.

## Reference

1. F. Wu, S. Zhao, B. Yu, et al., A new coronavirus associated with human respiratory disease in China, Nature 579, 7798, 265–269 (2020).

2. C. Huang, Y. Wang, et al., Clinical features of patients infected with 2019 novel coronavirus in Wuhan, China, Lancet 395, 10223, 497–506 (2020).

3. T. Singhal, A review of coronavirus disease-2019 (COVID-19), Indian J. Pediatr. 87, 281–286 (2020).

4. A. Padhi, S. Pradhan, P. P. Sahoo, K. Suresh, B. K. Behera, P. K. Panigrahi, Studying the effect of lockdown using epidemiological modelling of COVID-19 and a quantum computational approach using the Ising spin interaction, DOI: 10.1038/s41598-020-78652-0.

5. M. Pal, Genomic sequence data analysis using chaos game representation and mean structural similarity index measure to understand COVID-19 strains impacting wave 2 pandemic in India, DOI: 10.13140/RG.2.2.16342.78401.

6. M. Pal, A Novel integrative method for genomic sequence classification detecting mutant variants–A case study using the method applied to understand COVID-19 strains impacting wave 2 pandemic in India, DOI: 10.13140/RG.2.2.14379.16168.

7. J. P. Kanne, B.P. Little, J.H. Chung, B.M. Elicker, L.H. Ketai, Essentials for radiologists on COVID-19: an update—radiology scientific expert panel, Radiology (2020), DOI: 10.1148/radiol.2020200527.

8. H. X. Bai, B. Hsieh, et al., Performance of radiologists in differentiating COVID-19 from viral pneumonia on chest CT, Radiology (2020), DOI: 10.1148/radiol.2020200823.

9. X. Xie, Z. Zhong, W. Zhao, C. Zheng, F. Wang, J. Liu, Chest CT for typical 2019-nCoV pneumonia: relationship to negative RT-PCR testing, Radiology (2020), DOI: 10.1148/radiol.2020200343.

10. A. Schnuriger, M. Perrier, and et.al., Caution in interpretation of SARS-CoV-2 quantification based on RT-PCR cycle threshold value, DOI: 10.1016/j.diagmicrobio.2021.115366

11. D. Singh, V. Kumar, Vaishali, M. Kaur, Classification of covid-19 patients from chest ct images using multi-objective differential evolution–based convolutional neural networks, European Journal of Clinical Microbiology & Infectious Diseases, 1–11 (2020).

12. P. Rajpurkar, J. Irvin, et al., Chexnet: Radiologist-Level Pneumonia Detection on Chest X-Rays with Deep Learning, 2017 arXiv preprint 1711.05225.

13. E. E. D. Hemdan, M.A. Shouman, M.E. Karar, COVIDX-Net: A Framework of Deep Learning Classifiers to Diagnose COVID-19 in X-Ray Images, 2020 arXiv preprint 2003.11055.

14. L. Wang, Z. Q. Lin, A. Wong, COVID-Net: A Tailored Deep Convolutional Neural Network Design for Detection of COVID-19 Cases from Chest Radiography Images, 2020 arXiv preprint 2003.09871.

15. I. D. Apostolopoulos, T. Bessiana, COVID-19: Automatic Detection from X-Ray Images Utilizing Transfer Learning with Convolutional Neural Networks, 2003.11617.

16. A. Narin, C. Kaya, Z. Pamuk, Automatic Detection of Coronavirus Disease (COVID-19) Using X-Ray Images and Deep Convolutional Neural Networks, 2020 arXiv preprint 2003.10849.

17. Y. Song, S. Zheng, L. Li, X. Zhang, X. Zhang, Z. Huang, Y. Chong, Deep learning enables accurate diagnosis of novel coronavirus (COVID-19) with CT images, medRxiv (2020).

18. S. Wang, B. Kang, J. Ma, X. Zeng, M. Xiao, J. Guo, B. Xu, A deep learning algorithm using CT images to screen for Corona Virus Disease (COVID-19), medRxiv (2020).

19. C. Zheng, X. Deng, Q. Fu, Q. Zhou, J. Feng, H. Ma, X. Wang, Deep learning-based detection for COVID-19 from chest CT using weak label, medRxiv (2020), DOI: 10.1101/2020.03.12.20027185.

20. X. Xu, X. Jiang, C. Ma, P. Du, X. Li, S. Lv, et al., Deep Learning System to Screen Coronavirus Disease 2019 Pneumonia, 2020 arXiv preprint 200209334.

21. M. Barstugan, U. Ozkaya, S. Ozturk, Coronavirus (COVID-19) Classification Using CT Images by Machine Learning Methods, 2020 arXiv preprint 2003.09424.

22. E. Asnaoui, Y. Chawki, Using X-ray images and deep learning for automated detection of coronavirus disease, DOI: 10.1080/07391102.2020.1767212.

23. T. Ozturk, M. Talo, E. A. Yildirim, U. B. Baloglu, O. Yildirim, U. R. Acharya, Automated detection of COVID-19 cases using deep neural networks with X-ray images, Computers in Biology and Medicine 121, 103792 (2020).

24. Y. Pathak, P. K. Shukla, K.V. Arya, Deep bidirectional classification model for COVID-19 disease infected patients, DOI: 10.1109/TCBB.2020.3009859, IEEE/ACM

25. A. P. Adedigba, S. A. Adeshina, O. E. Aina, A. M. Aibinu, Optimal hyperparameter selection of deep learning models for COVID-19 chest X-ray classification; Intelligence-Based Medicine 5, 100034 (2021).

26. A. Z. Khuzani, M. Heidari, S. A. Shariati, COVID□Classifier: an automated machine learning model to assist in the diagnosis of COVID□19 infection in chest X□ray images; DOI: 10.1038/s41598-021-88807-2

27. P. K. Chaudhary, R. B. Pachori, FBSED based automatic diagnosis of COVID-19 using X-ray and CT images, DOI: 10.1016/j.compbiomed.2021.104454.

28. S. Pathan, P.C. Siddalingaswamy, T. Ali, Automated Detection of Covid-19 from Chest X-ray scans using an optimized CNN architecture, DOI: 10.1016/j.asoc.2021.107238

29. S. Toraman, T. B. Alakuş, İ. Türkoğlu, Convolutional CapsNet: A novel artificial neural network approach to detect COVID-19 disease from X-ray images using capsule networks, DOI: 10.1016/j.chaos.2020.110122

30. J. Li, G. Zhao, Y. Tao, P. Zhai, H. Chen, H. He, T. Cai, Multi-task contrastive learning for automatic CT and X-ray diagnosis of COVID-19, DOI: 10.1016/j.patcog.2021.107848

31. F. Demir, DeepCoroNet: A deep LSTM approach for automated detection of COVID-19 cases from chest X-ray images, DOI: 10.1016/j.asoc.2021.107160

32. M. Pal, P. K. Panigrahi, Effective clustering and accurate classification of the chest X-ray images of COVID-19 patients from healthy ones through the mean structural similarity index measure, DOI: 10.13140/RG.2.2.33801.57441

33. S. R. Nayak, D. R. Nayak, U. Sinha, V. Arora, R. B. Pachori, Application of deep learning techniques for detection of COVID-19 cases using chest X-ray images: A comprehensive study, DOI: 10.1016/j.bspc.2020.102365.

34. X. Mei, H. -C. Lee and et. al., Artificial intelligence–enabled rapid diagnosis of patients with COVID-19, DOI: 10.1038/s41591-020-0931-3

35. J. P. Cohen, COVID-19 Image Data Collection, 2020. https://github.com/ieee8023/COVID-chestxray-dataset.

36. X. Wang, Y. Peng, L. Lu, Z. Lu, M. Bagheri, R.M. Summers, Chestx-ray8: hospital scale chest x-ray database and benchmarks on weakly-supervised classification and localization of common thorax diseases, in: Proceedings of the IEEE Conference on Computer Vision and Pattern Recognition, 2097–2106 (2017).

37. G. Vaidelienė, J. Valantinas, The use of Haar wavelets in detecting and localizing texture defects, DOI: 105566/ias.1561.

38. J. Iacovacci, L. Lacasa, Visibility graphs for image processing, IEEE Transactions on Pattern Analysis and Machine Intelligence, DOI: 10.1109/TPAMI.2019.2891742.

39. L. Lacasa, J. Iacovacci, Visibility graphs of random scalar fields and spatial data, Phys. Rev. E 96, 012318 (2017).

40. L. Lacasa, B. Luque, F. Ballesteros, J. Luque, J. C. Nunoe, From time series to complex networks: The visibility graph, PNAS, DOI: 10.1073/pnas.0709247105.

41. D. Zhu, S. Semba, H. Yang, Matching Intensity for Image Visibility Graphs: A New Method to Extract Image Features, IEEE Access, DOI:10.1109/ACCESS.2021.3050747.

42. P. K. Sethy, S. K. Behera, Detection of Coronavirus Disease (COVID-19) Based on Deep Features, Preprints (2020) DOI:10.20944/preprints202003.0300.v1

43. M. Toğaçar, B. Ergen, Z. Cömert, COVID-19 detection using deep learning models to exploit social mimic optimization and structured chest X-ray images using fuzzy color and stacking approaches, Comput. Biol. Med. 103805 (2020).

44. F. Ucar, D. Korkmaz, COVIDiagnosis-Net: Deep Bayes-SqueezeNet based diagnosis of the coronavirus disease 2019 (COVID-19) from X-Ray images, Med. Hypotheses 140, 109761 (2020).

